# The potential economic impact of the updated COVID-19 mRNA fall 2023 vaccines in Japan

**DOI:** 10.1101/2023.12.04.23299402

**Authors:** K Fust, K Joshi, E Beck, M Maschio, M Kohli, A Lee, Y Hagiwara, N van de Velde, A Igarashi

**Affiliations:** Quadrant Health Economics Inc; 92 Cottonwood Crescent, Cambridge, Ontario, Canada; Moderna, Inc., 200 Technology Square, Cambridge, MA, 02139, USA; Moderna, Inc., 4-1-1 Toranomon, Kamiyacho, Minato ward, Tokyo, 105-6923, Japan; Graduate School of Pharmaceutical Sciences, University of Tokyo, 7-3-1 Hongo, Bunkyo City, Tokyo, 113-0033, Japan; Graduate School of Data Sciences, Yokohama City University School of Medicine, 22-2 Seto, Kanazawa Ward, Yokohama, Kanagawa Prefecture, 236-0027, Japan

## Abstract

This analysis estimates the economic and clinical impact of a Moderna updated COVID-19 mRNA Fall 2023 vaccine for adults ≥18 years in Japan. A previously developed Susceptible-Exposed-Infected-Recovered (SEIR) model with a 1-year analytic time horizon (September 2023-August 2024) and consequences decision tree were used to estimate symptomatic infections, COVID-19–related hospitalizations, deaths, quality-adjusted life-years (QALYs), costs, and incremental cost-effectiveness ratio (ICER) for a Moderna updated Fall 2023 vaccine versus no additional vaccination, and versus a Pfizer-BioNTech updated mRNA Fall 2023 vaccine. The Moderna vaccine is predicted to prevent 7.2 million symptomatic infections, 272,100 hospitalizations and 25,600 COVID-19 related deaths versus no vaccine. In the base case (healthcare perspective), the ICER was ¥1,300,000/QALY gained ($9,400 USD/QALY gained). Sensitivity analyses suggest results are most affected by COVID-19 incidence, initial vaccine effectiveness (VE), and VE waning against infection. Assuming the relative VE between both bivalent vaccines apply to updated Fall 2023 vaccines, the base case suggests the Moderna version will prevent an additional 1,100,000 symptomatic infections, 27,100 hospitalizations, and 2,600 deaths compared to the Pfizer-BioNTech vaccine. The updated Moderna vaccine is expected to be highly cost-effective at a ¥5 million willingness-to-pay threshold across a wide range of scenarios.

## Introduction

The first confirmed case of COVID-19 infection in Japan occurred on January 15, 2020.^1^ Since then, there have been over 33.8 million cases and almost 75,000 COVID-19 related deaths reported.^2^ COVID-19 vaccinations in Japan started on February 17, 2021, with priority to health care workers, before expanding to the broader population. On May 8, 2023, following the World Health Organization (WHO) announcement that the world pandemic was over,^3^ Japan changed the immunization category of COVID-19 from Category 2, which includes diseases such as tuberculosis, to Category 5, which is in the same classification as seasonal influenza.^4,5^ This meant that government funding for COVID-19 screening and treatment changed such that Spring 2023 COVID-19 vaccinations were only publicly funded for specific high-risk populations, whereas the Fall 2023 COVID-19 campaign covered the general population aged 6 months and above. The COVID-19 vaccine will continue to be funded and procured directly through the Japanese government until March 2024, but it is expected that the Japanese government will transition COVID-19 vaccines to the traditional National Immunization Program (NIP) in 2024 with regular NIP funding mechanisms from April 2024 onwards.^6^

First booster vaccinations in Japan became available in December 2021, where individuals were boosted primarily with one of the two available mRNA COVID-19 vaccines.^7,8^ Japanese studies have shown that vaccine effectiveness (VE) varies depending on the variant, as well as the version of vaccines being administered.^8–14^ Adapting to the changing variants, Moderna and Pfizer-BioNTech both updated their vaccines to a bivalent version against BA.4/BA.5 for Fall 2022. For Fall 2023, monovalent versions of the vaccine against the XBB.1.5 versions are available for administration. While the Moderna and Pfizer-BioNTech vaccines have the same mechanism of action, their formulations differ, for instance, in terms of lipid nanoparticles (delivery system) and dosage. Studies on previous versions of the mRNA vaccines have found higher VE values for different versions of the Moderna mRNA-1273 vaccine compared to the Pfizer-BioNTech BNT162b2 versions, including high-risk populations.^15–21^

Given the new Fall 2023 vaccine campaign, with updated Fall 2023 vaccines from both Moderna and Pfizer-BioNTech it is important to determine the economic and clinical consequences of vaccination during the Fall 2023, and understand the impact of newly emerging variants, to assist in the decision-making process and inform currently ongoing NIP discussions. Additionally, as the Japanese government has previously recommended and funded a Spring season vaccination campaign for select high-risk populations, the economic and clinical consequences of continuing to fund this population needs to be explored, as it likely will be a discussion subject for the NIP in 2024.

Therefore, the objective of this analysis is to estimate the economic and clinical impact of a NIP vaccination campaign from April 2024 onwards using the Fall 2023 updated mRNA COVID-19 Moderna vaccine compared to the Fall 2023 updated mRNA COVID-19 Pfizer-BioNTech vaccine for adults 18 years of age and older, using a previously developed Susceptible-Exposed-Infected-Recovered (SEIR) model.^22^ The impact of a Spring 2024 vaccination campaign in select, high-risk individuals (60-64 high-risk and 65+ general population; 65+ general population only) is also explored.

## Methods

### Overview

Two sets of comparisons were performed using a 1-year analytic time horizon (September 2023 to August 2024). First, vaccination of individuals aged ≥18 years with the updated Moderna COVID-19 mRNA Fall 2023 vaccine (Moderna Fall Campaign) was compared to no additional COVID-19 vaccination in Fall 2023 (No Fall Vaccine). Second, the Moderna Fall Campaign was compared to vaccination of individuals aged ≥18years with the updated Pfizer-BioNTech COVID-19 mRNA Fall 2023 vaccine (Pfizer-BioNTech Fall Campaign). A previously published^23^ SEIR model was used to estimate the total number of infections, and a decision tree was used to calculate infection-related consequences including numbers of symptomatic infections, COVID-19 related hospitalizations and deaths, treatment-related costs, and quality-adjusted life-years (QALYs) for the Moderna Fall Campaign, No Vaccine, and Pfizer-BioNTech Fall Campaign. The incremental cost effectiveness ratio (ICER), in terms of incremental cost per QALY gained, comparing the Moderna Fall Campaign and No Fall Vaccine was estimated. the economically justifiable price (EJP) differences between the Moderna and Pfizer-BioNTech vaccine was estimated at different willingness-to-pay thresholds (WTP) (¥5 million, ¥6 million, ¥10 million).^24,25^ The base-case was conducted from the healthcare cost perspective, while a scenario analysis was conducted using the societal cost perspective. Results are presented in both Japanese yen and United States dollars, using a conversion rate of ¥1 = 0.006996 USD.^26^

### SEIR Model

A previously developed SEIR model was used to calculate the number of infections (symptomatic and asymptomatic) as well as the VE against hospitalization for this analysis.^22,27^ This model has been adapted to Japan and has been used to project incidencefor the analytical time horizon of September 2023 to August 2024. The base case incidence projections and all of the incidence scenario analyses have been previously described.^28^ In the current analysis, we use all of the same inputs from the previous Japanese analysis except for VE as described below. As described in the previous analysis, we assume uptake of the Fall 2023 vaccine by age is similar to the first COVID-19 booster in Japan. ^28^ The VE against infection reduces the incidence of asymptomatic or symptomatic infection on each day of the simulation. The average incremental protection against hospitalization due to COVID-19 infection is also calculated on a monthly basis within the simulation and used within the infection consequences model. The number of infections predicted by the SEIR model is summarized by month for use in the infection consequences model. The proportion of infections with symptoms, assumed to be 67.6% based on a meta-analysis for Omicron variants,^29^ is applied as only symptomatic infections have economic consequences.

### Fall 2023 Vaccine Effectiveness Inputs

Japanese Vaccine Effectiveness Real-Time Surveillance for SARS-CoV-2 (VERSUS) is an ongoing test-negative case-control study examining the VE of the COVID-19 vaccines in individuals aged 16 years and over.^30^ Results are reported by different time frames corresponding to periods of different variant dominance, and with different versions of the mRNA vaccines (e.g. monovalent primary series, monovalent boosters, bivalent boosters).^8–14^ Neutralizing antibody titre data have shown a strong immune response from the Moderna Fall 2023 vaccine to XBB.1.5.^31^, however, data on clinical outcomes will not be available until 2024. For our previous Japanese analysis,^28^ we developed a base case and ranges that reflect the values reported across the different VERSUS reports.

For the base case of the current analysis, we assumed that the Moderna and Pfizer-BioNTech Fall 2023 vaccines are well-matched to the dominant circulating variant at the time. The VE values of the Moderna Fall 2023 vaccine are therefore predicted based on existing VE values of the bivalent vaccines against the BA.4/BA.5 variants from the VERSUS study.^32^ Although titre data may indicated a stronger response with the higher dose updated monovalent XXB.1.5 vaccine, data from the bivalent was deemed most appropriate because it is the most recently administered vaccine with real-world evidence data.^33^ The VE against infection and of those between 16-64 years old and those aged ≥65 years was 54.7% and 75.2% respectively. The model does not accommodate different VE inputs by age groups, so 54.7% was applied in the base case to all ages as the initial VE against infection, with the 95% confidence interval (40.3% - 65.6%) being used in sensitivity analyses. Due to sample size, the VE for hospitalizations was not disaggregated by age groups in this phase of the VERSUS study and was found to be 84.9% overall. This estimate was used for base case with the 95% confidence intervals (65.7% - 93.3%) applied in sensitivity analyses.

In order to approximate the VE against infection and hospitalization for the Pfizer-BioNTech Fall 2023 vaccine, the relative Ves (rVEs) between the Moderna and Pfizer-BioNTech bivalent vaccines for infections and hospitalizations were assumed to be maintained for the Fall 2023 vaccine. The rVE values for aged ≥ 18 years were obtained from a retrospective cohort study from the US, which estimated the adjusted rVE for hospitalizations and outpatient visits of the Moderna bivalent booster compared to the Pfizer-BioNTech bivalent booster using cox regression models. The rVE was defined as 100*(1 – adjusted hazard ratio from the analysis) and was estimated to be 5.1% (95% confidence interval: 3.2%-6.9%) for outpatient and 9.8% (95% confidence interval 2.6% - 16.4%) for hospitalization.^21^ As the rVE values for infection were not reported, the rVEs for outpatient visits were used as a proxy for infection. Final VE inputs are displayed in Table 1.

**Table 1.** Base case initial vaccine effectiveness values.

|  | **Infection** | **Hospitalization** |
| --- | --- | --- |
| **Moderna updated Fall 2023 vaccine** |  |  |
| **Base case\*** | 54.7% | 84.9% |
| **Scenario analysis compared to no vaccine: 95% confidence intervals** | 40.3% - 65.6% | 65.7% - 93.3% |
| **Pfizer-BioNTech updated Fall 2023 vaccine** |  |  |
| **Base case** | 52.3% | 83.3% |
| **Scenario analysis (Moderna vs. Pfizer-BioNTech): based on rVE 95% confidence interval (Moderna updated Fall 2023 vaccine initial VE held constant).** | 51.3%-53.2% | 81.9% - 84.5% |

\* These values were also used for the following scenarios: 1) Target population of 65 years and above; 2) Target population of 60 – 64 years (high risk) plus 65 years and above.

The monthly waning rates of both vaccines were based on data from a meta-analysis on the duration of protection from monovalent boosters against infection and hospitalization during the Omicron period.^34^ These values were 4.8% for infection, and 1.4% for hospitalization. In sensitivity analyses, the 95% confidence intervals were used: 3.1%-6.8% for infection and 0.6%-2.4% for hospitalization.

### Consequences Model Structure and Inputs

Monthly outputs from the SEIR model, including total number of symptomatic infections and incremental reduction in risk of hospitalization for vaccinated versus unvaccinated cohorts are used in a decision tree (Figure 1) to calculate the clinical and economic consequences of infections. Although not depicted in the decision tree figure, the model includes an infection-related myocarditis toll. The risk of infection-related myocarditis ^35^ is assumed to vary by age, and cost and QALY losses are applied to those patients experiencing infection-related myocarditis. Independent of the myocarditis risk, a proportion of COVID-19 infections are assumed to be treated in hospital, either in a general ward or more severe intensive care unit (ICU) setting. The age-dependent probabilities of hospitalization in the unvaccinated were derived previously.^28^ The percent of cases treated in the ICU care were calculated based on an insurance claims analysis of the DeSC database.^36^ Hospitalization probabilities are reduced according to VE for those who are vaccinated; VE is assumed to vary by vaccination strata and is calculated within the SEIR model. Death associated with COVID-19 is assumed to occur in hospitalized patients only (Table 2).^37,38^. Inputs are displayed in Table 2.

**Figure 1.**
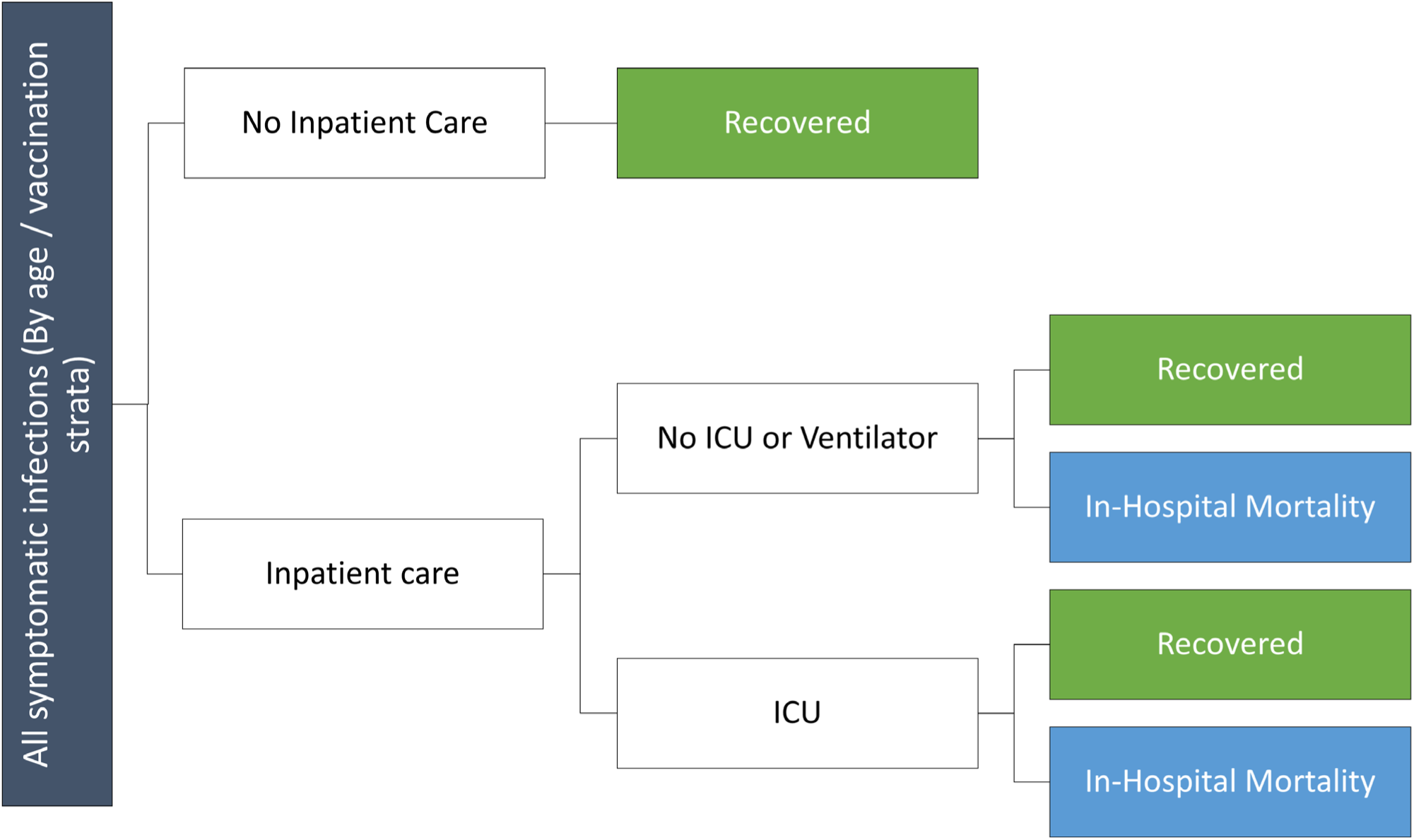
Consequences diagram.

**Table 2.** Base case probabilities for the economic consequences model.

| **Age Group (years)** | **Hospitalization in the unvaccinated<sup>28</sup>** | **Proportion hospitalized who receive ICU care<sup>36</sup>** | **In-hospital mortality<sup>37,38</sup>** | **COVID-19 infection–related myocarditis, by age (%)<sup>35</sup>** |
| --- | --- | --- | --- | --- |
| 0-11 | 0.46% | 17.35% | 0.14% | 0.12% |
| 12-17 | 0.18% | 17.35% | 0.14% | 0.12% |
| 18-29 | 0.30% | 33.21% | 0.15% | 0.08% |
| 30-39 | 0.47% | 36.39% | 0.15% | 0.07% |
| 40-49 | 0.51% | 51.39% | 1.10% | 0.09% |
| 50-59 | 1.04% | 51.39% | 1.10% | 0.14% |
| 60-64 | 2.93% | 56.38% | 4.98% | 0.14% |
| 65-74 | 8.62% | 52.56% | 7.07% | 0.16% |
| 75+ | 13.76% | 55.85% | 12.91% | 0.21% |

ICU: Intensive care unit

For the healthcare cost perspective, the average cost of outpatient care was calculated from an insurance claims analysis of the DeSC database, which derives information from one insurance system for retired persons and one designed for persons aged 75 years and above in Japan.^36^ Costs in the claims analysis were estimated by establishing a baseline period of 1 to 3 months prior to the onset of COVID-19 infection and comparing the incremental medical costs for the 6 months following COVID-19 infection to baseline for the following age groups: 0-19 years, 20-39 years, 40-59 years, 60-64 years, 65-74 years, 75-84 years, and ≥85 years.^36^ Accordingly, outpatient costs utilized in the base-case analyses reflect a weighted average (calculated using the Japanese population distribution across age groups) of the 6-month COVID-19 attributable cost. The cost of outpatient care was weighted by the probability of seeking outpatient care for treatment to account for the percentage of patients with non-severe COVID-19 who do not seek medical attention. A similar approach was employed for hospitalization costs, which were also estimated using insurance claims from the DeSC database and represent the 6-month COVID-19 attributable cost.^36^ As both the outpatient and hospitalization costs reflect the 6-month period following infection, explicit post-infection and long COVID costs are not included as they are assumed to be reflected in the base-case cost inputs. The infection-related myocarditis cost was estimated to be ¥20,262 based on the DeSC data. Given the 1-year time horizon for the analytic period, none of the costs were discounted. Base case cost inputs are displayed in Table 2.

The expected number of life years lost due to early deaths from COVID-19 was calculated using expected survival by age as reported by e-Stat (Official Statistics of Japan).^39^ Age-specific utility values for individuals without infection, obtained from Shiroiwa (2021), were attached to each year lost due to early death from COVID-19.^40^ All future QALYs lost were discounted by 2% annually to present value.^41^ QALY losses associated with COVID-19 infection were estimated based on EQ-5D survey data measured up to 90 days following infection onset from a Japanese clinic for patients treated whether in-hospital or on an outpatient basis using an area under the curve approach.^42^ QALY losses for hospitalized patients also include 3 days of outpatient impact to account for the period of time prior to hospitalization with infection symptoms. As base-case QALY losses reflect the 3-month period following infection, post-infection and long COVID QALY loss impacts are assumed to be reflected in the base-case inputs and are not explicitly included. Baseline utility estimates and QALY losses associated with infection-related outcomes are presented in Tables 3 below.

### Vaccine-Related Costs and QALYs Lost

Vaccine costs included an administration cost of ¥3,400^43^ and the unit cost of the vaccine itself. As the commercial price of the vaccines in Japan has not yet been determined, a unit cost of both vaccines was assumed to be ¥12,040 in the base case based on the German list price.^44^ All individuals receiving vaccines also received an average cost and QALY loss associated with adverse events (AE), weighted by the probability of experiencing the AE. AEs include local and systemic grade 3 reactions, anaphylaxis, and vaccine-induced myocarditis (relevant for those ages 18-39 years only); risks were estimated based on Moderna clinical trial data and published sources.^45–47^ The unit cost of myocarditis following COVID-19 infection was assumed to apply to the vaccine-induced myocarditis AE.^48^ All other AE costs were derived from Teng (2022), with the assumptions that 20% of grade 3 systemic AEs would require an outpatient visit, and that 60% of anaphylaxis cases would require hospitalization (and the remaining 40% would require emergency room care).^49^ AE QALY losses were obtained from Teng (2022), and the QALY loss associated with infection-induced myocarditis was estimated based on data from Prosser (2019).^49,50^ Inputs for vaccine-related AEs are presented in Table 4 below.

**Table 3.** Base case cost and quality of life inputs for the economic consequences model.

| **Model Parameter** | **Value** |
| --- | --- |
| Discount rate<sup>41</sup> | 2.0% |
| **Costs** |  |
| Booster cost (per dose) | ¥12,040 |
| Booster administration cost<sup>43</sup> | ¥3,400 |
| COVID-19 infection–related myocarditis (per event)<sup>48</sup> | ¥20,262 |
| Outpatient care (per patient seeking care)*<sup>36</sup> | ¥65,361 |
| Hospitalization (no ICU)* <sup>36</sup> | ¥508,683 |
| Hospitalization (ICU)* <sup>36</sup> | ¥1,366,787 |
| **Quality of Life** |  |
| Baseline utility data<sup>40</sup> |  |
| 0-11 years | 0.973 |
| 12-17 years | 0.973 |
| 18-29 years | 0.955 |
| 30-39 years | 0.949 |
| 40-49 years | 0.946 |
| 50-59 years | 0.928 |
| 60-64 years | 0.928 |
| 65-74 years | 0.929 |
| 75+ years | 0.832 |
| QALYs lost, not hospitalized<sup>42</sup> | 0.0367 |
| QALYs lost, hospitalized<sup>† 42</sup> | 0.05 |
| QALYs lost, SARS-CoV-2 infection–related myocarditis (per event)<sup>50</sup> | 0.0019 |

\* Reflects the excess COVID-attributable cost for the 6-month period following infection (including acute treatment)
<sup>†</sup>Includes an additional 3 days of disutility to account for the symptomatic time prior to hospitalization

**Table 4.**
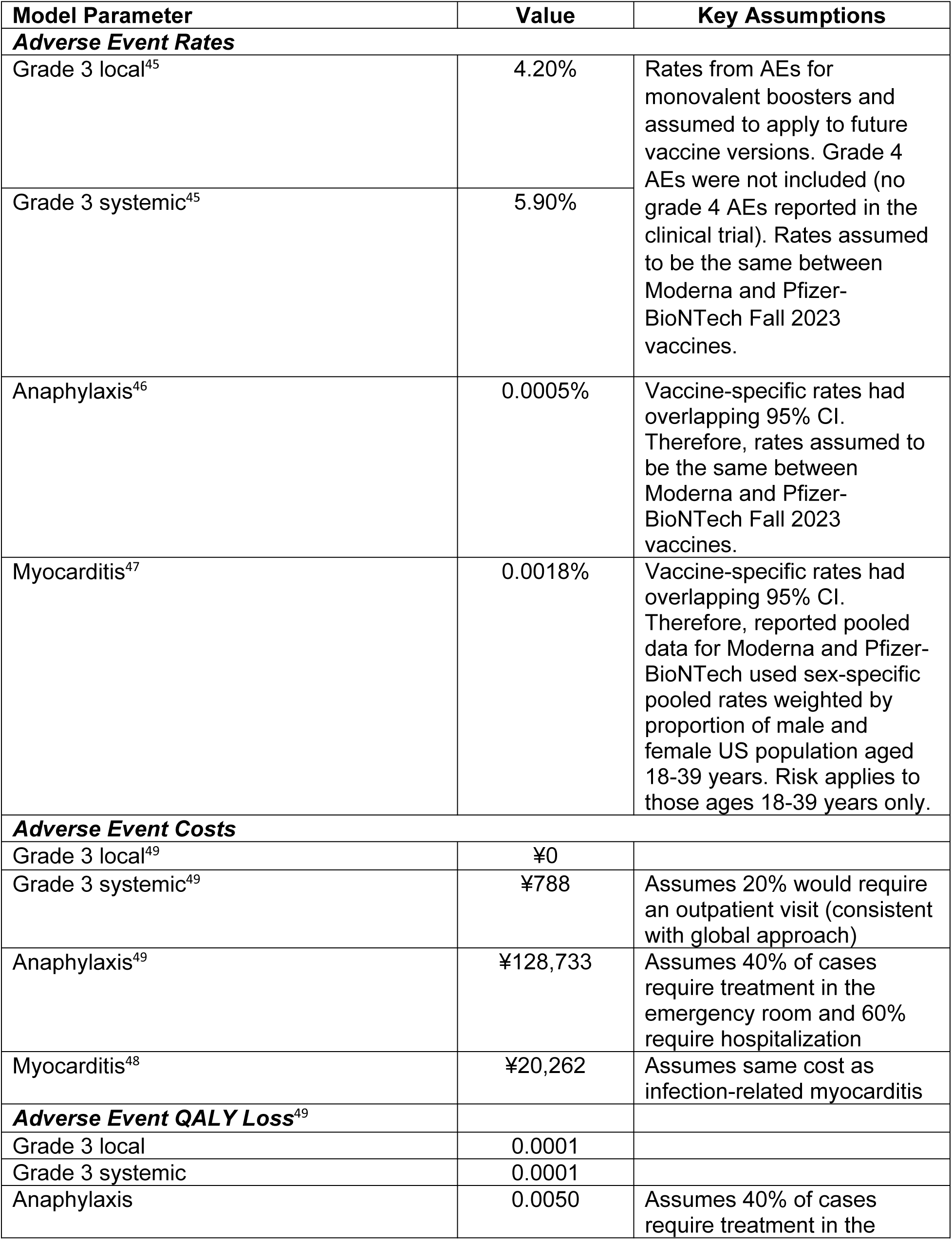
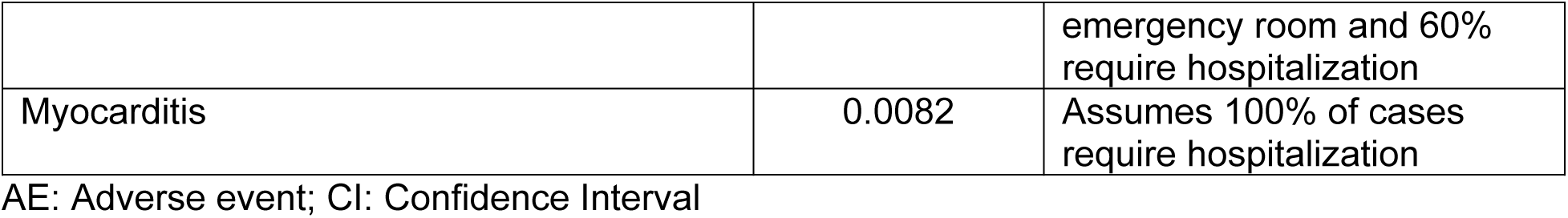
Adverse event inputs.

### Analysis of Uncertainty

A number of scenario analyses and deterministic sensitivity analyses were conducted to determine the impact of uncertainty on the predicted ICER of the Moderna Fall Campaign compared to No Fall Vaccine. The discount rate applied to the calculation of lifetime QALYs was varied from 0% to 4% as per the Japanese guidelines.^41^ The societal cost perspective was also considered using the inputs displayed in Table 5. Days lost from work associated with vaccination and vaccine-related AEs, as well as infection-related consequences, were applied but weighted by age based on the proportion of individuals in the workforce.^51^ Days of lost productivity were valued assuming an average daily wage of ¥18,700.^52^

**Table 5.** Lost productivity for analysis from the societal perspective.

| **Model Parameter** | **Value** | **Key Assumptions** |
| --- | --- | --- |
| Percentage in labor force<sup>51</sup> |  |  |
| 0-11 years | 0.0% |  |
| 12-17 years | 23.3% |  |
| 18-29 years | 63.2% |  |
| 30-39 years | 86.4% |  |
| 40-49 years | 86.4% |  |
| 50-59 years | 82.3% |  |
| 60-64 years | 78.1% |  |
| 65-74 years | 25.2% |  |
| 75+ years | 25.2% |  |
| Daily wage rate<sup>52</sup> | ¥18,700 |  |
| Days lost for: |  |  |
| Vaccination | 0.6 | Assumption |
| Infection, not hospitalized<sup>64</sup> | 5 |  |
| Infection, hospitalized* | 15 | Includes time loss for the hospitalization length of stay (10 days) and an additional 5 days to reflect time with symptoms prior to hospitalization |
| SARS-CoV-2 infection–related myocarditis | 3 | Assumption based on US data |
| Grade 3 Local AE | 0.50 | Assumption |
| Grade 3 Systemic AE | 0.50 | Assumption |
| Myocarditis<sup>65</sup> | 2.25 | Assumption based on US data (inpatient length of stay of 2.25 days and 8 hours missed per day) |
| Anaphylaxis<sup>65</sup> | 2.00 | Assumption based on US data (inpatient length of stay of 2 days and 8 hours missed per day) |

Alternative vaccination target populations for the Fall 2023 Campaign were tested as Japan has varied the population eligible for booster doses over time. A subgroup analysis was conducted where vaccines were offered only to those aged 65 and older. A second analysis included those aged 65 years and older plus those aged 60 to 64 years at high risk of severe outcomes. We estimated the number of high-risk individuals ages 60-64 to be 4.48 million, or 60%, through an analysis of the DeSC data.^36^

One scenario was created to estimate the impact of an annual strategy with two doses. As in the base case, the first dose was provided as Fall 2023 vaccination to those 18 and older starting in September. A second dose was then assumed to be provided to those 18 – 64 at high risk of severe outcomes and those 65 years of age and older starting 6 months later in March 2024. The coverage rate for this second vaccine was assumed to mirror the observed bivalent dose which had similar eligibility criteria. The technical appendix of Kohli et al. estimating the corresponding clinical impact provides the uptake over time.^28^

The inputs to the SEIR model were varied in several scenarios. The cost-effectiveness of vaccination was tested using the six different incidence strategies that were developed in Kohli et al.^28^ These scenarios were created by changing the calibration process (Adjusted Tokyo data 2.5X or 1.5X; Adjusted Tokyo data 2.0x with Revised Waning), a scenario assuming change in contact patterns during the year (Seasonality: phi=0.2), and two further scenarios assuming that a new variant that reduced vaccine VE and natural immunity evolved (immune escape assumed to happen either April 2024 or June 2024). Please see the previous clinical manuscript for further details.^28^ Vaccine coverage was also reduced in scenarios to 50% and 75% of base case values. The initial VE of the Moderna vaccine and the monthly waning rates were varied as described in the vaccine section above.

Deterministic sensitivity analyses were conducted by varying inputs in the infection consequences decision tree. Percent with symptoms, mortality rates, and percent in the ICU were varied using the lower and upper bounds of the 95% confidence interval associated with the base case input. Hospitalization costs, outpatient costs, and QALY losses were varied by +/- 25%. Finally, alternative prices for the Moderna vaccine were assumed to be ¥10,072 and ¥18,525, corresponding to the list price of $129.50 per dose in the United States, were tested.^53^

In comparing the Moderna and Pfizer-BioNTech Fall Campaign, the initial VE against both infection and hospitalization of the Pfizer-BioNTech Campaign was varied in sensitivity analyses using the values displayed in Table 1.

## Results

### Base Case Comparison: Updated Moderna Fall Campaign versus No Fall Vaccine

For the 18 year old plus base-case scenario, the model predicted 35,241,000 symptomatic infections without a fall vaccine, this decreased by 20% to 28,055,300 with the Moderna Fall Campaign (Table 6). The model predicted 690,000 COVID-19 related hospitalizations without a fall vaccine, compared to 417,800 with the Moderna Fall Campaign, a reduction of 39%. Model predicted deaths were also decreased from 62,000 to 36,100 by implementing the Moderna Fall Campaign (41% reduction). Base-case clinical results, which align with the previously published clinical manuscript, are presented in Table 6.^28^

**Table 6.** Base-case (18+ general population): Number of cases, hospitalizations, and deaths.

|  | Total |  |  | Prevented by Moderna Vaccine (% Decrease) |  |
| --- | --- | --- | --- | --- | --- |
|  | No Vaccine | Pfizer-BioNTech Vaccine | Moderna Vaccine | Vs. No Vaccine | Vs. Pfizer-BioNTech Vaccine |
| Symptomatic infections | 35,240,923 | 29,146,363 | 28,055,308 | 7,185,614 (20%) | 1,091,054 (4%) |
| Hospitalizations | 689,973 | 444,948 | 417,839 | 272,133 (39%) | 27,108 (6%) |
| Deaths | 61,738 | 38,736 | 36,128 | 25,610 (41%) | 2,607 (7%) |

Given the clinical impact of the updated Moderna COVID-19 mRNA Fall 2023 vaccine, it is expected to result in a gain of 207,000 QALYs relative to No Fall Vaccine by preventing COVID-19 related deaths and 266,800 QALYs gained due to prevented morbidity for a total of 473,900 QALYs gained. Vaccination and adverse events cost ¥1,123,500 million ($7,860 million USD) compared with savings in COVID-19 treatment costs of ¥485,900 million ($9,400 million USD) due to vaccination. The incremental cost per QALY gained of the updated Moderna COVID-19 mRNA Fall 2023 vaccine compared to no additional COVID-19 vaccination in Fall 2023 is therefore ¥1,300,000 ($9,400 USD) (Table 7; See Technical Appendix Table 1 – 2 for details).

**Table 7.**
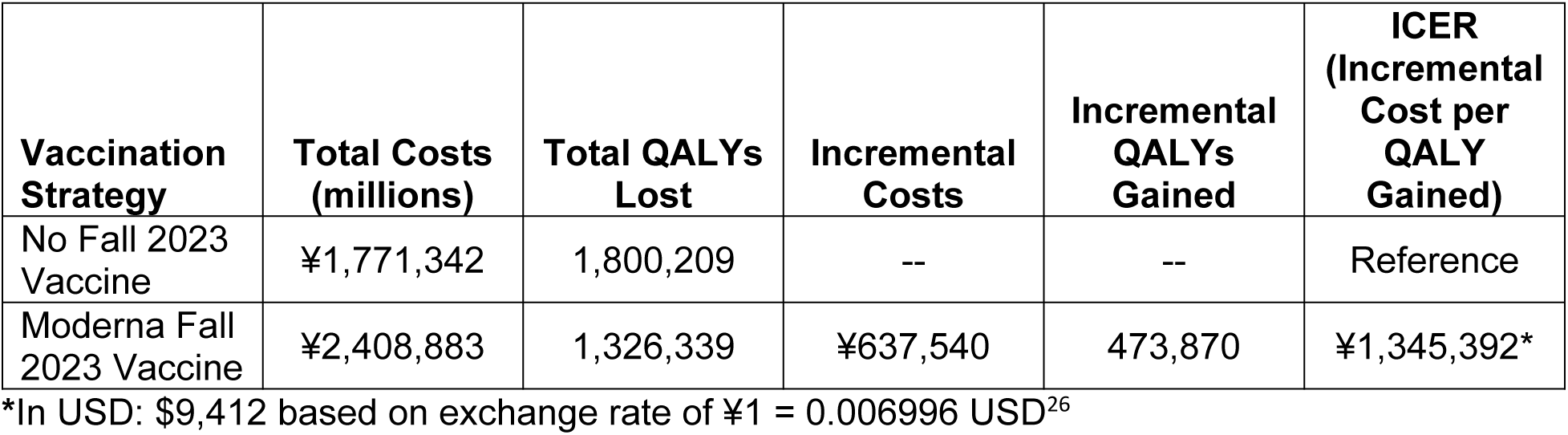
Base-case (18+ general population): Cost-Effectiveness Results.

| Vaccination Strategy | Total Costs (millions) | Total QALYs Lost | Incremental Costs | Incremental QALYs Gained | ICER (Incremental Cost per QALY Gained) |
| --- | --- | --- | --- | --- | --- |
| No Fall 2023 Vaccine | ¥1,771,342 | 1,800,209 | -- | -- | Reference |
| Moderna Fall 2023 Vaccine | ¥2,408,883 | 1,326,339 | ¥637,540 | 473,870 | ¥1,345,392* |

\*In USD: \$9,412 based on exchange rate of ¥1 = 0.006996 USD<sup>26</sup>

### Scenario Analysis: Societal Perspective and Target Population

Analyses performed from the societal cost perspective yield an ICER of ¥800,000/QALY gained ($5,600 USD/QALY gained), representing a decrease from the base-case ICER of 40%. Limiting the population vaccinated to high-risk individuals aged 60-64 years and those aged ≥65 years decreases the ICER compared to no Fall vaccine by 32% to ¥910,000 ($6,400 USD) compared to the base case (healthcare payer cost perspective). Limiting the population vaccinated to only those aged ≥65 years yields a similar ICER of ¥940,000/QALY gained ($6,600 USD/QALY gained).

### Scenario Analysis: Two Annual Boosters

In the scenario analysis examining a two-booster strategy, the total number of COVID-19 infections prevented by the Moderna Fall Vaccine Campaign compared to the no Fall Vaccine strategy increases by 28%, from the base-case value of 7,185,614 to 9,691,565. Incremental QALYs gained also increased from the base-case from 473,870 to 605,737. With the increased number of vaccinations and associated costs of providing a larger number of doses, total costs of the Moderna Vaccine Campaign also increase in the 2-booster scenario to ¥2,740,270 million ($19,171 million USD). As a result, the ICER increases from the base-case of ¥1,300,000 ($9,400 USD) to ¥1,600,000 ($11,200 USD) per QALY gained relative to no vaccine.

### Additional Sensitivity Analyses

Varying the updated Moderna COVID-19 mRNA Fall 2023 vaccine price yields ICERs of ¥1,000,000 ($7,300 USD) per QALY gained and ¥2,300,000 ($16,400) per QALY gained relative to no fall vaccine, representing a decrease of 22% and increase of 74% respectively. Results of the remaining sensitivity analyses are displayed in Figure 2, with economic details in Technical Appendix Table 3 and clinical details in Technical Appendix Table 4. COVID-19 incidence, vaccine waning against infection, and initial VE against infection have the greatest impact on model results due to their effect on the number of infections prevented by vaccination. For example, with incidence scenario, “Adjusted Tokyo Data (2.5X)”, the timing of the incidence of cases has changes so that a larger proportion of infections occur before the vaccine is administered. With this scenario, the ICER increases by 318% to ¥5,600,000 per QALY ($39,000 USD), as the Fall 2023 vaccine is predicted to prevent 80% fewer infections relative to the base-case (1,465,400 compared to 7,185,600). Varying vaccine waning rates against infection has the next largest impact, yielding ICERs ranging from ¥600,000 ($4,200 USD) to ¥4,300,000 ($30,000 USD) per QALY. This is also due to the large impact on COVID-19 infections prevented (1,464,382 [80%] for the infection waning rate lower bound and 12,077,866 [168%] for the upper bound). Similarly, initial VE against infection has the third strongest impact on the ICER, which ranges from ¥540,000 ($3,800 USD) to ¥3,800,000 ($26,900) per QALY gained relative to no vaccine using the upper and lower bounds, respectively. Using the lower bound infection waning rate yields 1,756,555 infections prevented, while varying to the upper bound yields 12,648,644 infections prevented, representing a 76% decrease and 176% increase from the base case, respectively. The pattern is similar for the other incidence scenarios (adjusted Tokyo data [2.0x] revised waning, adjusted Tokyo data [1.5x], immune escape June 2024, and immune escape April 2024), as well as the percentage of infection with symptoms, which appear toward the top of the tornado diagram (Figure 2) due to their impact on infections prevented by vaccination.

**Figure 2.**
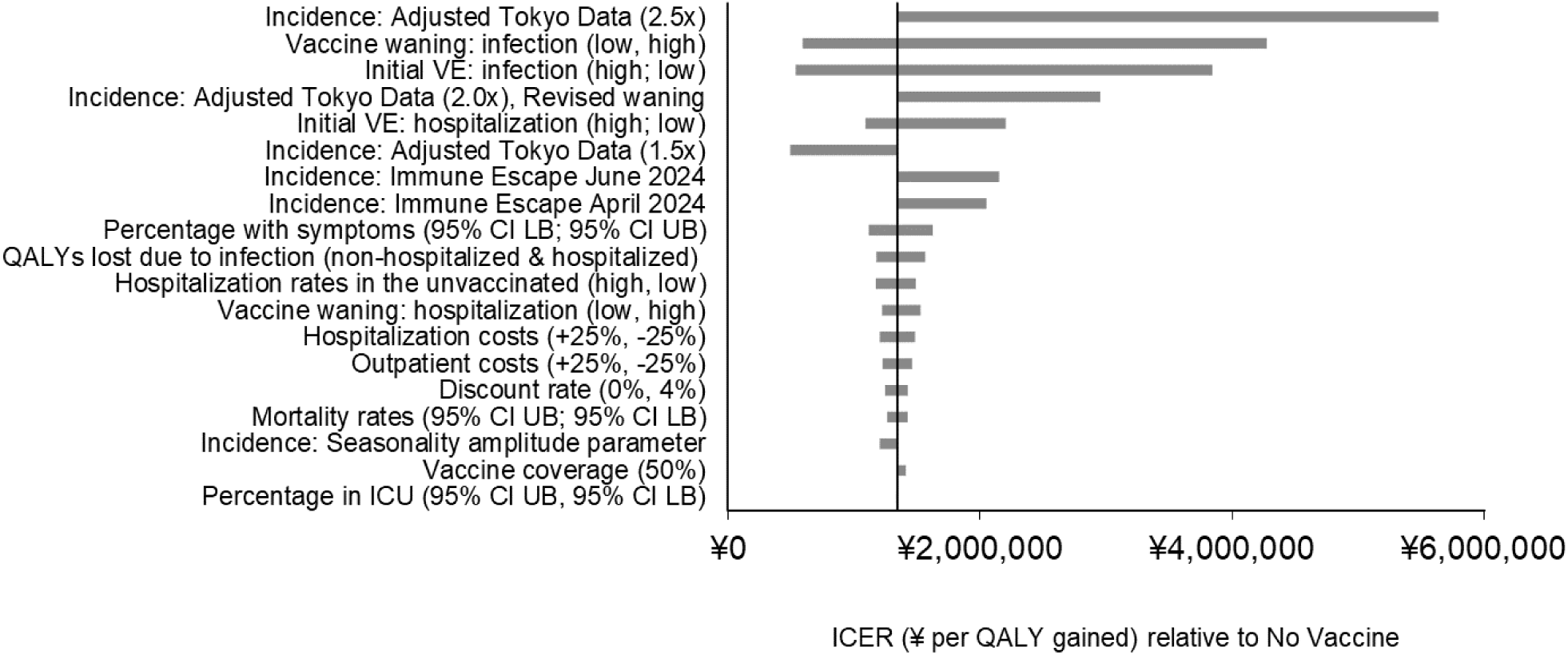
Impact of scenario analyses on the projected incremental cost-effectiveness results with a COVID-19 vaccine updated for Fall 2023.

Initial VE against hospitalization also has a substantial impact on the ICER, with variation from the upper and lower bounds yielding ICERs of ¥1,100,00 ($7,600 USD) and ¥2,200,00 ($15,000 USD) per QALY gained relative to no vaccine. These results are driven by the total number of hospitalizations prevented which range from 338,397 to 120,673 respectively compared to the base case estimate of 272,133. QALY losses due to infection (both hospitalized and non-hospitalized varied simultaneously) yield ICERs ranging from ¥1,200,000 to ¥1,600,000 per QALY gained relative to no vaccine. Hospitalization rates in the unvaccinated, vaccine waning against hospitalization, and hospitalization costs appear together in the tornado diagram, due to their similar impact on number of hospitalizations prevented by vaccination and importance of hospitalization costs in driving cost-effectiveness results. All other parameters varied in deterministic sensitivity analyses yield changes from the base case ICER of less than 10%.

### Comparison: Moderna Fall Campaign versus Pfizer Fall Campaign

The predicted initial VE of the updated Moderna Fall 2023 vaccine was based on input data assumed to be greater than the initial VE of the updated Pfizer-BioNTech version, leading to a reduction in the total number of symptomatic infections of 1,091,100 (4%). The Moderna Fall Campaign is also predicted to reduce the total number of hospitalizations by 6%, and COVID-19 related deaths by 7%. These reductions in clinical outcomes translate to economic benefits, with the Moderna Fall Campaign leading to a gain of 60,800 QALYs (4% increase) and COVID-19 treatment cost savings of ¥60,400 million ($422 million USD; 5% reduction compared to the Pfizer-BioNTech Fall ampaign). If the Moderna and Pfizer-BioNTech Fall vaccines are priced equivalently, the updated Moderna COVID-19 mRNA Fall 2023 vaccine would be considered the dominant choice.

In analyses comparing the Moderna Fall Campaign to the Pfizer-BioNTech Fall Campaign, the EJP difference for the updated Moderna COVID-19 mRNA Fall 2023 vaccine is presented in Technical Appendix Table 5. Given the assumption of superior effectiveness of the updated Moderna COVID-19 mRNA Fall 2023 vaccine, a higher price for the Moderna vaccine relative to the Pfizer-BioNTech vaccine could be justified economically. Assuming a base-case price per dose of ¥12,040 ($84 USD), the price difference at a ¥5 million WTP threshold is ¥5,022 ($35 USD), while the price differences at the ¥6 and ¥10 million WTP thresholds are ¥5,860 ($41 USD) and ¥9,212 ($64 USD), respectively. As differences between the Moderna and Pfizer-BioNTech VE increase or decrease, the price difference that could be justified at any WTP threshold also increases or decreases. For example, scenario analyses varying the relative VE to the upper and lower bounds suggest that the EJP difference could range from ¥2,829 ($20 USD) using the lower bound at a WTP threshold of ¥5 million up to ¥13,709 ($96 USD) using the upper bound at a WTP threshold of ¥10 million (Technical Appendix Table 5).

## Discussion

Using a previously published SEIR model^28^, this analysis examined the potential cost-effectiveness of an updated Moderna COVID-19 mRNA Fall 2023 vaccine administered to adults in Japan aged ≥18 years relative to no vaccine. An updated Moderna COVID-19 mRNA Fall 2023 vaccine is predicted to prevent 7.2 million symptomatic infections, 272,100 hospitalizations and 25,600 COVID-19 related deaths in Japan between September 2023 and August 2024 compared to no vaccination. In the base case analysis, the incremental cost per QALY gained was predicted to be ¥1,300,000 ($9,400 USD) using the healthcare payer cost perspective. Considering a WTP threshold of ¥5 million, the vaccine would be highly cost-effective if priced at ¥12,040. Considering a societal cost perspective yields an ICER of ¥800,000 per QALY gained. Changing the target population to vaccination of high-risk individuals aged 60-64 years combined with the general population aged 65+ or only the general population aged 65+ yields ICERs of ¥910,000 and ¥940,000 per QALY, respectively.

Overall, sensitivity analyses suggest that results are robust to parameter uncertainty and demonstrate that the cost-effectiveness model results are most affected by COVID-19 infection incidence, initial VE against infection, and VE waning against infection. These key drivers were also important to the clinical impact of vaccination in Japan in the clinical analysis by Kohli et al. that focused on the SEIR model adaptation process.^28^ Infection incidence and VE have a large impact on the number of infections prevented by vaccination, which in turn impacts the economic results. These results are also consistent with a previously published analysis performed examining the cost-effectiveness of vaccination in the United States (US), although ^23^ differences between results for the two jurisdictions exist. For example, varying the hospitalization rates in the unvaccinated appears to have a greater impact on the ICER in the US when compared to Japan. The range used in the US sensitivity analysis was wider than the range used for Japan. It incorporated uncertainty around the US hospitalization rates based on data from the Delta period and around the relative risk used to adjust those rates for Omicron. A limitation of the current analysis is that while hospitalization rates in Japan reflect data from April 2022 to March 2023, the vaccination status of hospitalized patients was not tracked. Hospitalization rates in the unvaccinated population were therefore estimated using by varying the assumptions about the level of residual VE due to prior vaccinations in the population.^28^

QALY losses associated with COVID-19 infection also appear to have a greater impact on model results in Japan compared to the US. This is likely due to differences in how QALY losses were estimated between the two countries. In Japan, estimates were based on EQ-5D survey data measured up to 90 days following infection onset from the VERSUS clinic for patients treated whether in-hospital or on an outpatient basis and represent a 3-month time period following infection onset^36^, while in the US^23^, separate QALY loss estimates were used for the acute and post-infection periods. Hospitalization costs appear to have a similar impact in both settings; however, outpatient costs appear to have a greater impact on model results in Japan compared to the US. In Japan, hospitalization and outpatient costs reflect the 6-month period following infection, while in the United States, separate cost estimates were used to reflect the acute and the 6 month post-infection periods.^23^ Furthermore, the QALY and cost impacts of the post-infection consequences of COVID-19 infections are likely under-estimated in this analyses as data is now showing the effects of “long COVID” are likely to last longer than 6 months.

The Centers for Disease Control and Prevention (CDC) in the United States has presented evidence that COVID-19 may impact quality-of-life for up to 2 years post-acute infection.^54–56^ In the United States, a database analysis was conducted on individuals that tested positive for COVID-19 between March – December 2020 and matched controls.^57^ Individuals were followed for 2 years to estimate the risk of pre-specified COVID-19 post-acute sequalae. The analysis found that at 2 years, 65% of individuals that were hospitalized for COVID-19 and 31% of those that were not hospitalized still had post-sequalae risks. These risks contributed to disability-adjusted life-year decrements (DALYs) in this population, with 25% and 21% of DALYs lost during the second year in non-hospitalized and hospitalized individuals, respectively. Based on data from Japan, our model includes 3 months of QALY decrements. Given the evidence that QALY loss extends beyond the 3-month period, and given fewer COVID-19 infections and cases of long COVID are estimated with the use of the Moderna Fall 2023 vaccine compared to the Pfizer-BioNTech Fall 2023 vaccine, our estimates of QALYs gained with the Moderna vaccine are likely underestimated.

A recently published analysis conducted in the United Kingdom (UK) using electronic health records from January 2020-January 2023 compared resource utilization in individuals with and without long COVID post-infection to controls.^58^ The resource use of the long COVID group was also compared to pre-2020 resource use. The study found that per year, individuals with long COVID had more general practitioner consultations and outpatient visits than all other control groups but fewer inpatient, critical care, and emergency department visits than those with COVID-19 without a long COVID diagnosis. Annual healthcare utilization cost for the long COVID group was estimated at £3,335 per year; the cost for those with COVID but not long COVID was £5,961 while the control groups had costs ranging from £1,210 to £1,283. Although healthcare utilization patterns and costs differ between the UK and Japan, this study demonstrates that long COVID costs extends beyond the 6 months included in our analysis. Additionally, even if not diagnosed with long COVID, healthcare utilization is increased in COVID patients compared to those that did not become infected. Similar to QALYs gained, our estimates of cost-savings with the Moderna Fall 2023 vaccine is likely underestimated.

This analysis also demonstrated the potential economic impact of a difference in vaccine effectiveness between the two Fall 2023 COVID-19 mRNA vaccines. Assuming that the rVE observed with the bivalent vaccine will also apply to the updated fall 2023 version, the base case comparison of the two mRNA vaccines indicates that the Moderna version will prevent 1,100,000 more symptomatic infections, 27,100 more hospitalizations, and 2,600 more deaths relative to the Pfizer-BioNTech vaccine. Therefore, the unit cost of the Moderna vaccine could be higher than the Pfizer-BioNTech vaccine. Under base-case assumptions, a price difference of ¥5,022 ($35 USD) would be economically justifiable considering a ¥5 million WTP threshold. Scenario analyses varying the VE suggest the EJP difference may range from ¥2,800 ($20 USD) to ¥7,500 ($52 USD) at the ¥5 million WTP threshold. While both of the mRNA COVID-19 vaccines have the same mechanisms of action, differences in VE may exist because their delivery systems and dosage differ. A meta-analysis in immunocompromised populations concluded that the Moderna version of the COVID-19 vaccine is more effective than the Pfizer-BioNTech version^59^ In the general population, several studies of the original monovalent versions make the same conclusion.^15–18^ For this analysis, we use evidence from a study that compares the bivalent versions^60^ in the general population.

Although scenario analyses have been included in the current analysis to explore the impact of incidence and VE on cost-effectiveness results, as noted in previous publications, the future incidence of infection, expected pattern of infection, and VE against infection and hospitalization remain highly uncertain and may impact the value of vaccination in the future.^23,28,61^ In addition to long-COVID inputs which may underestimate the cost savings and QALYs gained with the use of the Moderna Fall 2023 vaccine, due to lack of data, other parameters, which may increase the value of vaccination, were not included. Estimates of mortality included in the current analysis represent in-hospital mortality only, and therefore any COVID-19 related deaths occurring post-discharge are not captured. For the societal cost perspective, we considered short-term lost productivity but did not have data on long-term losses or other informal health care resource used, such as caregiver time, ^62^ or other impact of family spillover.^63^ The impact of COVID-19 using the societal cost perspective is therefore under-estimated.

Despite the limitations, our model consistently predicted that the Moderna updated COVID-19 mRNA Fall 2023 vaccine is highly cost-effective at a ¥5 million WTP threshold across a wide range of parameter values and scenario analyses. The VE of the new vaccines will not be known until after the vaccine is delivered and real world, such as VERSUS study, have completed their assessment. However, if VE is similar to past versions of the vaccine, then the updated Moderna mRNA Fall 2023 vaccine is expected to prevent a significant number of COVID-19 symptomatic infections, hospitalizations, deaths and associated health care costs both in comparison against no vaccination and in comparison against the Pfizer-BioNTech vaccine. Finally, a higher price for a more effective vaccine is economically justifiable. If the Moderna version of the Fall 2023 vaccine is more effective that the Pfizer-BioNTech vaccine, our model predicted that a higher unit cost is justifiable given the additional cases of symptomatic infections, hospitalizations and deaths.

## Supporting information

Technical Appendix

## Data Availability

All data produced in the present work are contained in the manuscript

