## Supplementary material for "The potential economic impact of the updated COVID-19 mRNA fall 2023 vaccines in Japan": Technical Appendix

**ADDITIONAL RESULTS TABLES**

**Table 1. Base-case Quality-Adjusted Life-Years (QALYs) Lost**

|  | **Total QALYs Lost** | | | **Total QALYs Gained (Moderna Vaccine Vs.)** | | **% Change (Moderna Vaccine Vs.)** | |
| --- | --- | --- | --- | --- | --- | --- | --- |
|  | **No Vaccine** | **Moderna Vaccine** | **Pfizer Vaccine** | **No Vaccine** | **Pfizer Vaccine** | **No Vaccine** | **Pfizer Vaccine** |
| Morbidity | 1,303,544 | 1,036,697 | 1,077,133 | 266,847 | 40,436 | 20.5% | 3.8% |
| Mortality | 496,664 | 289,642 | 309,998 | 207,022 | 20,356 | 41.7% | 6.6% |
| **Total** | **1,800,209** | **1,326,339** | **1,387,131** | **473,870** | **60,792** | **26.3%** | **4.4%** |

**Table 2. Base-case Disaggregated Economic Results**

| **Economic Results (Yen, in millions)** | | | | | | | |
| --- | --- | --- | --- | --- | --- | --- | --- |
|  | **Total Costs** | | | **Cost Difference**  **(Moderna Vaccine Vs.)** | | **% Change**  **(Moderna Vaccine Vs.)** | |
| **Costs:** | **No Vaccine** | **Moderna Vaccine** | **Pfizer Vaccine** | **No Vaccine** | **Pfizer Vaccine** | **No Vaccine** | **Pfizer Vaccine** |
| Vaccination* | ¥0 | ¥1,120,028 | ¥1,120,028 | ¥1,120,028 | ¥0 | -- | 0% |
| Adverse Events | ¥0 | ¥3,422 | ¥3,422 | ¥3,422 | ¥0 | -- | 0% |
| Outpatient Care | ¥1,106,588 | ¥885,165 | ¥919,240 | -¥221,423 | -¥34,076 | -20% | -4% |
| Hospitalization | ¥663,846 | ¥399,576 | ¥425,841 | -¥264,270 | -¥26,265 | -40% | -6% |
| Infection-Related Myocarditis | ¥908 | ¥692 | ¥722 | -¥216 | -¥30 | -24% | -4% |
| **Total (treatment-related)** | **¥1,771,342** | **¥1,285,433** | **¥1,345,803** | **-¥485,909** | **-¥60,370** | **-27%** | **-4%** |
| **Total**  **(healthcare perspective)** | **¥1,771,342** | **¥2,408,883** | **¥2,469,253** | **¥637,540** | **-¥60,370** | **36%** | **-2%** |
| Lost productivity | ¥1,852,273 | ¥1,595,264 | -- | -¥257,009 | -- | -14% | -- |
| **Total (societal perspective)** | **¥3,623,615** | **¥4,004,147** | **--** | **¥380,532** | **--** | **11%** | **--** |

**Table 3. Deterministic sensitivity analysis economic results (Moderna Updated Fall 2023 Vaccine relative to No Fall 2023 Vaccine)**

|  |  | **ICER**  **(¥ per QALY Gained)** | | **Change from**  **Base-Case** | | **Change from**  **Base-Case (%)** | |
| --- | --- | --- | --- | --- | --- | --- | --- |
| **Model Parameter** | **Variation** | **Low Value** | **High Value** | **Low Value** | **High Value** | **Low Value** | **High Value** |
| Moderna Fall 2023 Vaccine Cost | ¥10,072 - ¥18,525 | ¥1,044,141 | ¥2,338,205 | -¥301,251 | ¥992,813 | -22% | 74% |
| Discount rate | (0%, 4%) per guidelines | ¥1,247,145 | ¥1,428,575 | -¥98,246 | ¥83,183 | -7% | 6% |
| Perspective | Societal | ¥803,030 | | -¥542,362 | | -40% | |
| Target population | 60-64 high-risk and 65+ general | ¥911,015 | | -¥434,377 | | -32% | |
| Target population | 65+ general | ¥942,104 | | -¥403,288 | | -30% | |
| Vaccination strategy | 2 booster | ¥1,599,586 | | ¥254,194 | | 19% | |
| Hospitalization rates | See clinical manuscript | ¥1,491,297 | ¥1,173,984 | ¥145,905 | -¥171,408 | 11% | -13% |
| Mortality rates | 95% CI | ¥1,427,202 | ¥1,264,468 | ¥81,810 | -¥80,924 | 6% | -6% |
| Initial VE: hospitalization | Lower and upper bounds from VERSUS | ¥2,204,076 | ¥1,092,674 | ¥858,684 | -¥252,718 | 64% | -19% |
| Initial VE: infection | Lower and upper bounds from VERSUS | ¥3,843,331 | ¥539,116 | ¥2,497,939 | -¥806,276 | 186% | -60% |
| Vaccine waning: hospitalization | Decreased and increased fall 2023 waning | ¥1,223,119 | ¥1,529,712 | -¥122,273 | ¥184,320 | -9% | 14% |
| Vaccine waning: infection | Decreased and increased fall 2023 waning | ¥595,046 | ¥4,274,696 | -¥750,346 | ¥2,929,304 | -56% | 218% |
| Vaccine coverage | 50%, 75% | ¥1,411,804 | ¥1,397,851 | ¥66,412 | ¥52,459 | 5% | 4% |
| Incidence | Immune Escape April 2024 | ¥2,051,738 | | ¥706,346 | | 53% | |
| Incidence | Immune Escape June 2024 | ¥2,151,514 | | ¥806,122 | | 60% | |
| Incidence | Adjusted Tokyo Data (2.5x) | ¥5,634,508 | | ¥4,289,117 | | 319% | |
| Incidence | Adjusted Tokyo Data (1.5x) | ¥495,259 | | -¥850,133 | | -63% | |
| Incidence | Seasonality: phi=0.2 | ¥1,205,428 | | -¥139,964 | | -10% | |
| Incidence | Adjusted Tokyo Data (2.0x), Revised Waning | ¥2,954,554 | | ¥1,609,162 | | 120% | |
| Hospitalization cost | +25%, -25% | ¥1,205,970 | ¥1,484,813 | -¥139,421 | ¥139,421 | -10% | 10% |
| Percentage with symptoms | 95% CI (upper bound, lower bound) | ¥1,118,259 | ¥1,625,364 | -¥227,133 | ¥279,972 | -17% | 21% |
| Outpatient cost | +25%, -25% | ¥1,228,576 | ¥1,462,208 | -¥116,816 | ¥116,816 | -9% | 9% |
| QALY Losses (non-hospitalized, hospitalized) | +25%, -25% | ¥1,178,971 | ¥1,566,518 | -¥166,421 | ¥221,126 | -12% | 16% |
| Percentage in ICU | 95% CI (upper bound, lower bound) | ¥1,336,406 | ¥1,354,394 | -¥8,986 | ¥9,002 | -1% | 1% |

A sensitivity analyses was conducted with the unit price of the Moderna updated COVID-19 mRNA Fall 2023 vaccine unit cost. If the unit cost of the vaccine is increased from ¥12,040 to ¥32,513, the incremental cost-effectiveness ratio (ICER) for the Moderna vaccine compared to no vaccine will be ¥5 million per quality-adjusted life-years (QALYs) gained. If the unit cost is increased to ¥39,045 or ¥65,174 the cost per QALY gained will be ¥6 million and ¥10 million respectively.

**Table 4. Deterministic sensitivity analysis clinical results (Moderna Updated Fall 2023 Vaccine relative to No Fall 2023 Vaccine)**

|  | **Symptomatic infections** | | | **Hospitalizations** | | | **Deaths** | | |
| --- | --- | --- | --- | --- | --- | --- | --- | --- | --- |
| Scenario | No Vaccine | With Vaccine | Prevented | No Vaccine | With Vaccine | Prevented | No Vaccine | With Vaccine | Prevented |
| Base case | 35,240,923 | 28,055,308 | 7,185,614 | 689,973 | 417,839 | 272,133 | 61,738 | 36,128 | 25,610 |
| 60-64 high-risk and 65+ general | 35,240,923 | 32,077,805 | 3,163,118 | 689,973 | 484,803 | 205,170 | 61,738 | 40,427 | 21,311 |
| 65+ general | 35,240,923 | 32,375,370 | 2,865,553 | 689,973 | 510,882 | 179,090 | 61,738 | 42,404 | 19,335 |
| 2 booster* | 35,240,923 | 25,549,267 | 9,691,656 | 689,973 | 365,258 | 324,715 | 61,738 | 30,961 | 30,777 |
| Higher hospitalization rates | 35,240,923 | 28,055,308 | 7,185,614 | 799,370 | 483,249 | 316,121 | 71,558 | 41,868 | 29,690 |
| Lower hospitalization rates | 35,240,923 | 28,055,308 | 7,185,614 | 607,756 | 368,653 | 239,103 | 54,320 | 31,792 | 22,528 |
| Higher mortality rate | 35,240,923 | 28,055,308 | 7,185,614 | 689,973 | 417,839 | 272,133 | 68,072 | 39,831 | 28,241 |
| Lower mortality rate | 35,240,923 | 28,055,308 | 7,185,614 | 689,973 | 417,839 | 272,133 | 55,920 | 32,734 | 23,186 |
| Low hospital initial VE | 35,240,923 | 28,055,308 | 7,185,614 | 689,973 | 569,299 | 120,673 | 61,738 | 51,058 | 10,680 |
| High hospital initial VE | 35,240,923 | 28,055,308 | 7,185,614 | 689,973 | 351,576 | 338,397 | 61,738 | 29,597 | 32,141 |
| Low infection initial VE | 35,240,923 | 33,484,368 | 1,756,555 | 689,973 | 482,140 | 207,833 | 61,738 | 42,242 | 19,496 |
| High infection initial VE | 35,240,923 | 22,582,279 | 12,658,644 | 689,973 | 343,660 | 346,313 | 61,738 | 29,291 | 32,447 |
| Low waning (infection) | 35,240,923 | 23,163,057 | 12,077,866 | 689,973 | 350,508 | 339,465 | 61,738 | 29,973 | 31,765 |
| High waning (infection) | 35,240,923 | 33,776,540 | 1,464,382 | 689,973 | 492,991 | 196,982 | 61,738 | 43,129 | 18,609 |
| Low waning (hospitalization) | 35,240,923 | 28,055,308 | 7,185,614 | 689,973 | 387,503 | 302,470 | 61,738 | 33,137 | 28,601 |
| High waning (hospitalization) | 35,240,923 | 28,055,308 | 7,185,614 | 689,973 | 458,419 | 231,554 | 61,738 | 40,130 | 21,608 |
| Vaccine Coverage (75% of base case) | 35,240,923 | 30,083,501 | 5,157,422 | 689,973 | 486,125 | 203,848 | 61,738 | 42,449 | 19,289 |
| Vaccine Coverage (50% of base case) | 35,240,923 | 31,859,918 | 3,381,005 | 689,973 | 553,622 | 136,350 | 61,738 | 48,771 | 12,968 |
| Immune Escape April 2024 | 43,148,725 | 38,087,118 | 5,061,607 | 842,573 | 612,108 | 230,465 | 74,951 | 53,768 | 21,183 |
| Immune Escape June 2024 | 39,068,442 | 34,279,489 | 4,788,953 | 759,749 | 532,840 | 226,909 | 67,622 | 46,392 | 21,230 |
| Adjusted Tokyo Data (2.5x) | 33,985,577 | 32,520,032 | 1,465,545 | 566,310 | 420,523 | 145,788 | 51,468 | 37,864 | 13,604 |
| Adjusted Tokyo Data (1.5x) | 33,606,420 | 22,216,834 | 11,389,586 | 829,881 | 408,036 | 421,845 | 72,775 | 34,126 | 38,648 |
| Seasonality: phi=0.2 | 36,426,098 | 28,859,505 | 7,566,593 | 723,775 | 432,012 | 291,762 | 65,096 | 37,601 | 27,496 |
| Adjusted Tokyo Data (2.0x), Revised Waning* | 39,927,629 | 38,219,091 | 1,708,538 | 888,266 | 612,731 | 275,534 | 78,996 | 53,710 | 25,286 |
| Low percentage with symptoms | 31,534,370 | 25,104,521 | 6,429,849 | 617,403 | 373,892 | 243,511 | 55,245 | 32,329 | 22,916 |
| High percentage with symptoms | 38,942,262 | 31,001,946 | 7,940,316 | 762,440 | 461,725 | 300,715 | 68,222 | 39,923 | 28,299 |

*** 2 Booster Strategy: One dose is offered to adults aged 18 years and older in Fall 2023; A second booster dose is offered to high risk individuals aged 18 – 64 and all individuals ages 65 years and older in Spring 2024.**

**Table 5. Economically Justifiable Price Difference* (Moderna Price Difference vs. Pfizer-BioNTech)**

|  | **Moderna Price Difference vs. Pfizer-BioNTech^†^** | | |
| --- | --- | --- | --- |
| **Scenario** | **¥5 mil WTP** | **¥6 mil WTP** | **¥10 mil WTP** |
| Base-case | ¥5,022 | ¥5,860 | ¥9,212 |
| Relative vaccine effectiveness lower bound | ¥2,829 | ¥3,303 | ¥5,199 |
| Relative vaccine effectiveness upper bound | ¥7,480 | ¥8,726 | ¥13,709 |

^†^Vaccine administration cost is equivalent for Moderna and Pfizer vaccines, so estimates reflect the economically justifiable price difference

* The price difference between Moderna and Pfizer-BioNTech is the same regardless of the unit cost of the Moderna vaccine.
